# The performance of national COVID-19 ‘Symptom Checkers’: A comparative case simulation study

**DOI:** 10.1101/2020.04.28.20084079

**Authors:** F Mansab, S Bhatti, D Goyal

## Abstract

**Introduction:** The response to COVID-19 differs from nation to nation. There are likely a number of factors one can attribute to such disparity, not least of which is differing healthcare models and approaches. Here, we examine the COVID-19 community triage pathways employed by four nations, specifically comparing the safety and efficacy of national online ‘symptom checkers’ utilised within the triage pathway.

**Methods:** A simulation study was conducted on current, nationwide, patient-led symptom checkers from four countries (Singapore, Japan, USA and UK). 52 cases were simulated to approximate typical COVID-19 presentations (mild, moderate, severe and critical), and COVID-19 mimickers (e.g. sepsis and bacterial pneumonia). The same simulations were applied to each of the four country’s symptom checkers, and the recommendations to refer on for medical care or to stay home were recorded and compared.

**Results:** The symptom checkers from Singapore and Japan advised onward healthcare contact for the majority of simulations (88% and 77% respectively). The USA and UK symptom checkers triaged 38% and 44% of cases to healthcare contact, respectively. Both the US and UK symptom checkers consistently failed to identify severe COVID-19, bacterial pneumonia and sepsis, triaging such cases to stay home.

**Conclusion:** Our results suggest that whilst ‘symptom checkers’ may be of use to the healthcare COVID-19 response, there is the potential for such patient-led assessment tools to worsen outcomes by delaying appropriate clinical assessment. The key features of the well performing symptom checkers are discussed.

**SUMMARY:** *What is already known?:*

- The availability and use of symptom checkers are increasing.
- Symptom checkers are currently in use at a national level to help in the healthcare response to COVID-19.
- There is limited evidence to support the effectiveness or safety of symptom checkers as triage tools during a pandemic response.

*What does this paper add?:*

- This study compares performance of symptom checkers across different countries, revealing marked variation between national symptom checkers.
- The symptom checkers employed by Japan and Singapore are twice as likely to triage cases onward for clinical assessment than those of the US or UK.
- The US and UK symptom checkers frequently triaged simulated cases of sepsis, bacterial pneumonia and severe COVID-19 to stay home with no further healthcare contact.
- We discuss the key aspects of the well-performing triage systems.

## INTRODUCTION

COVID-19 is a new infection in humans. The symptom profile, disease progression and complication rates are still relatively unknown [1]. From the available evidence, four broad categories of illness have been postulated. ‘Mild COVID-19’ makes up over 80% of cases, and is typically a self-limiting infection similar to the common cold, resolving without intervention. ‘Moderate COVID-19’ typically has features of viral pneumonia in the absence of hypoxia, progressing to ‘Severe COVID-19’ typically when patients require oxygen therapy. ‘Critical COVID-19’, where ventilatory support is typically required, occurs in less than 5% of cases [2]. The rate of disease progression is not fixed: early intervention and various management strategies can reduce the rate of progression to critical illness and death [2-6].

Whilst the infection fatality rate (IFR) is yet to be determined, COVID-19 is associated with a substantive mortality. Over a period of five months COVID-19 has led to more than 300,000 deaths; with more than half these deaths occurring within the last month [7].

The risk of mortality is affected by a number of risk factors. Co-existing health problems such as diabetes, heart disease and cancer have been implicated as conferring a higher risk of mortality in COVID-19 [8]. Age appears to be the most striking and consistent risk factor for COVID-related mortality [9]. Based on current data, the mortality rate in patients under 50 years of age is thought to be less than 1.1%, rising to around 14% in those over 80 years of age [10].

Variation in mortality also seems to exist between countries [11]. Initially this variation was thought to be predominantly related to the method of recording deaths and the total number of tests conducted (i.e., the detection of milder cases) [12]. As the pandemic spreads across the globe, it is becoming increasingly clear that how a country responds to the pandemic impacts the number of deaths their locality will experience [6,11].

The national response to the COVID-19 pandemic has many important tenants. On the public health side, infection control initiatives attempt, in part, to mitigate the surge of infections that can accompany new pathogens where there is little circulating immunity. This reduces mortality by preventing the healthcare services from being overwhelmed, thus permitting improved access to medical management for those who need it [6]. The clinical response to COVID-19 also centres on access to treatment. To successfully reduce the mortality rate, those patients who are developing more severe disease must be identified [3].

Identifying those COVID-19 patients that require treatment is challenging. Firstly, COVID-19 has a broad range of presentations that can mimic common conditions that rarely require clinical assessment (e.g., the common cold) [1]. Secondly, there are no clinical signs or symptoms that reliably predict who will progress to severe disease [3]. As such, the clinical community is left with a large number of potential cases without any clear symptom indicators for 1) who has the disease, and 2) who is developing more severe disease. The problem is compounded further as more serious, life-threatening conditions (e.g., bacterial pneumonia and sepsis) can mimic any stage of COVID-19 disease [13,14].

National ‘Symptom Checkers’ have been implemented in many countries in the hope of reducing this burden faced by healthcare services. Symptom-checkers are self-assessment tools. The individual - typically online or via computer application - enters their symptoms into a predetermined platform, and from there a predetermined algorithm produces an outcome (usually advice). This is a form of self-led triage. It is hoped that such self-directed assessments will enable the identification of potential cases [15], and will correctly triage those individuals who would benefit from clinical assessment and/or management into further care [16]. For such a hope to be realised, symptom checkers must be able to determine mild conditions from severe conditions.

Whilst self-triage has been used for some years in non-emergency conditions to varying degrees of success [17], self-triage has never before been used in a pandemic setting, and as yet the efficacy and safety has not be formally studied. Caution must be exercised, as, to date, studies examining symptom checkers have had mixed and disappointing results in general - demonstrating poor diagnostic performance (34-58%) and questionable triage performance (55-80%) [18]. The stakes are high, in that a failure to triage serious medical conditions (such as Severe COVID-19, bacterial pneumonia or sepsis) in for further assessment will inevitably lead to delayed treatment and higher mortality [19-22].

Here, we test the performance of four nationwide symptom checkers from four nations to ascertain how safe and efficient each symptom checker is in differentiating mild from severe COVID-19 cases, and how well they detect time-sensitive COVID-19 mimickers such as bacterial pneumonia and sepsis.

## METHODOLOGY

Five countries were initially selected for analysis. Three (Singapore, Japan and Norway) were selected as they maintained low case fatality rates despite a demonstrable surge of cases in the preceding two months. Two countries (the UK and the USA) were selected due to concern regarding high case fatality rates.

Public Health guidelines from each country were reviewed. Access was obtained to any available government sponsored online patient-led triage system (Singapore-‘Singapore COVID-19 Symptom Checker’ - [23], Japan - “Stop COVID Symptom Checker” - [24], USA - ‘CDC Coronavirus Symptom Checker’ - [25] and the UK - ‘111 COVID Symptom Checker’- [26]). Whereas the NHS ‘111’ COVID-19 Symptom Checker was, and continues to be heavily utilised (with over 500,000 assessments completed on average each month [27]), there was no available data as to the usage of the other symptom checkers.

For the purpose of this analysis, data was extracted only from those countries with symptom checkers (Singapore, Japan, UK and USA), in an effort to compare the performance of symptom checkers specifically.

### Case Scenario’s

52 standardised cases were designed simulating common COVID-19 related presentations with varying severity or risk factors.

Case scenarios included four distinct presentations: (1) Cough and fever; (2) Co-morbidity, cough and fever; (3) Immunosuppression, cough and fever, and (4) Shortness of Breath and fever. These distinct presentations were then varied in relation to one or more of the following (1) Duration of symptoms; (2) Age of Patient, and (3) Severity of symptoms.

‘Fever’ was chosen as a core symptom of COVID-19 due to its high discriminatory value for infection. Even though it may only be present in less than half of COVID-19 cases at presentation [28], the presence of fever permits greater focus on infective causes in relation to shortness of breath and cough. Fever also presents commonly in sepsis and pneumonia [29]; two of the key diagnoses that triage systems need to detect to prevent excess mortality. Fever has also been shown to relate to disease severity and mortality outcomes in COVID-19 [30].

‘Cough’ is a non-specific symptom covering a wide range of conditions. Combined with fever, cough raises the possibility of chest infection, including COVID-19 and bacterial pneumonia (one of the critical differential diagnosis’s in COVID-19). Detecting possible bacterial pneumonia is a pre-requisite to a functioning triage system given the time-critical manner to antibiotic initiation to prevent unnecessary deaths [30].

‘Shortness of breath’ is generally accepted as a marker of COVID-19 disease progression [31], albeit there are other reasons for shortness of breath, and specifically in COVID-19, patients may not experience shortness of breath despite being hypoxic - so called, ‘silent hypoxia’ [32]

‘Duration’ was chosen as a severity marker as the prolongation of fever, cough and/or shortness of breath within the context of COVID-19 or a COVID-19 mimicker (pneumonia, sepsis, etc…) carries a worse prognosis. In particular, an unremitting, persistent fever warrants further assessment in regard to COVID-19 [30], but also in relation to sepsis [29].

‘Age’ is a well defined risk factor for severe complications of COVID-19 [9,10]. As such, it was deemed useful to include age as a variable in the case simulations to test whether the symptom checker accounted for age when determining risk.

‘Severity’ of symptoms relates to duration of fever, cough and shortness of breath. Shortness of breath had its own severity scale, and was crucial for staging level of complicated COVID-19, severity of pneumonia and sepsis [29,30]. Mild Shortness of Breath was defined as shortness of breath during activities that did not stop one completing the activity. Moderate shortness of breath was defined differently depending on age. That is, respiratory reserve was considered to be less in adults > 70 yrs of age in comparison to the younger age groups, and as such we defined moderate shortness of breath in those > 70 yrs of age as preventing the completion of most tasks, whilst for younger cases moderate shortness of breath would still permit most tasks to be completed. Severe shortness of breath was defined as shortness of breath at rest.

The immunosuppression case simulations related to the development of cough and fever four days after chemotherapy, simulating potential neutropenic sepsis. Neutropenic sepsis is a medical emergency requiring immediate medical attention, and early antibiotic therapy - door to needle time for sepsis should be less than one hour, and for neutropenic sepsis less than 30 minutes [33,34].

Except for the paediatric case, hypertension was chosen as the co-morbidity due to its discriminatory value between mild and severe co-morbidities. There is evidence that hypertension may be an independent risk factor for poorer outcomes in COVID-19, however it remains, as do many of the proposed ‘high-risk’ co-morbidities, unproven [8]. Differentiating symptom checkers that account for milder co-morbidities or make allowances for the uncertainty that remains in the evidence base for at risk groups was deemed useful in regard to symptom checkers’ safety performance.

Where equivocal answers existed, such as for breathless: ‘yes’, ‘I’m not sure’ or ‘no’, the equivocal answer (‘I’m not sure’) was interpreted as mild symptoms. Unless stated in the specific case scenario, any question pertaining to co-morbidity was answered as ‘no’. All other variations were as described for each case scenario (Supplementary Data).

### Statistical Analysis

The primary outcome was total number of cases referred onward for further clinical assessment which was converted into a percentage ratio and then compared between countries.

## RESULTS

The key baseline population and testing data are presented in Table 1. Notably, the highest rate of testing for COVID-19 was by Singapore with the lowest being Japan.The UK had the highest reported physicians per capita, Japan and Singapore had the lowest. Cases per thousand inhabitants varied greatly, with Singapore and the UK maintaining similar rates. From the available statistics Singapore had the lowest CFR (<0.1%) and the UK had the highest CFR (13.6%) currently. All population and testing data was extracted from The WHO as of 26.04.2020.

**Table 1.** Key population and COVID-19 testing data from each of the five countries.

|  | Singapore | Japan | USA | UK |
| --- | --- | --- | --- | --- |
| **Population Data** |  |  |  |  |
| Total Tests (per million) | 20,815 | 1,166 | 16,507 | 9,867 |
| Total Tests (thousands) | 122 | 147 | 5,500 | 669 |
| Population (millions) | 5.8 | 126.5 | 331 | 67.9 |
| Confirmed COVID-19 Positive | 12,693 | 13,182 | 899,281 | 148,381 |
| Cases per thousand inhabitants | 2.2 | 0.1 | 2.7 | 2.3 |
| Case Fatality Rate (%) | <0.1 | 2.7 | 5.6 | 13.6 |
| Physicians per 10,000 head of capita | 24 | 24 | 25 | 28 |

52 case scenarios were applied to each country’s patient-led triage systems. The results for each scenario are presented in tabulated format (Supplementary Data). Singapore had the highest overall referral rate at 88%, and the US had the lowest at 38% (table 2).

**Table 2.** Total number (percentage) of case simulations referred on by country. Distinct scenarios are included. Variation within each scenario is not detailed here (See Supplementary Data).

|  | Case Fatality Rate % | Total cases referred onwards<br>N=52 (%) | Cough + Fever<br>N=16 (%) | Comorbidity + Cough + Fever<br>N=12 (%) | Immunosuppressed + Cough + Fever<br>N=12 (%) | Shortness of Breath + Fever<br>N=12 (%) |
| --- | --- | --- | --- | --- | --- | --- |
| Singapore | 0.1 | 46 (88) | 10 (63) | 12 (100) | 12 (100) | 12 (100) |
| Japan | 2.7 | 40 (77) | 12 (75) | 8 (67) | 8 (75) | 12 (100) |
| US | 5.6 | 20 (38) | 0 | 3 (25) | 12 (100) | 5 (42) |
| UK | 13.6 | 23 (44) | 0 | 3 (25) | 12 (100) | 8 (67) |

From the cases not referred, the US and UK triaged a significant number of cases to ‘stay home’ that would normally have required early intervention or urgent care. The US triage system (CDC Coronavirus Symptom Checker) frequently triaged home case simulations with possible severe COVID-19, bacterial pneumonia and sepsis, and triaged possible neutropenic sepsis to healthcare contact within 24hours. The UK ‘111’ COVID-19 self-triage system, frequently triaged possible severe COVID-19 and bacterial pneumonia to stay at home with no follow-up, and is likely to have delayed treatment for sepsis, severe COVID-19 and neutropenic sepsis. It is of note that whilst Japan’s symptom checker generally performed well, our simulation revealed a potential delay to treatment for neutropenic sepsis. Indeed, all four symptom checkers failed to triage the simulation for neutropenic sepsis into the “emergency department”. (Table 3).

**Table 3.** Tabulated view of likely triage outcome of specific diagnosis in each country. Columns indicate clinical diagnosis and rows represent the likely consequence of the country’s triage response. Red indicates cases that would have likely been dismissed (stay home) by the patient-led triage system. Orange indicates cases that were likely to have been triaged to delayed clinical contact, or to stay at home. Green indicates diagnoses likely to have been captured and triaged to clinical care. URTI - Upper Respiratory Tract Infection

|  | URTI (mild) | COVID (mild) | COVID (moderate) | Bacterial Pneumonia | Sepsis | COVID (severe) | Neutropenic Sepsis | Septic Shock | COVID (Critical) |
| --- | --- | --- | --- | --- | --- | --- | --- | --- | --- |
| USA | Red | Red | Red | Red | Red | Orange | Orange | Orange | Green |
| UK | Red | Red | Red | Red | Orange | Orange | Orange | Green | Green |
| Japan | Red | Orange | Green | Green | Green | Green | Orange | Green | Green |
| Singapore | Red | Orange | Green | Green | Green | Green | Orange | Green | Green |

### High CFR versus Low CFR Countries

The main differences in triage criteria extrapolated from the national symptom checkers relating to COVID-19 between the low case fatality rate (CFR) countries and the high CFR countries are presented at Table 4.

**Table 4.**
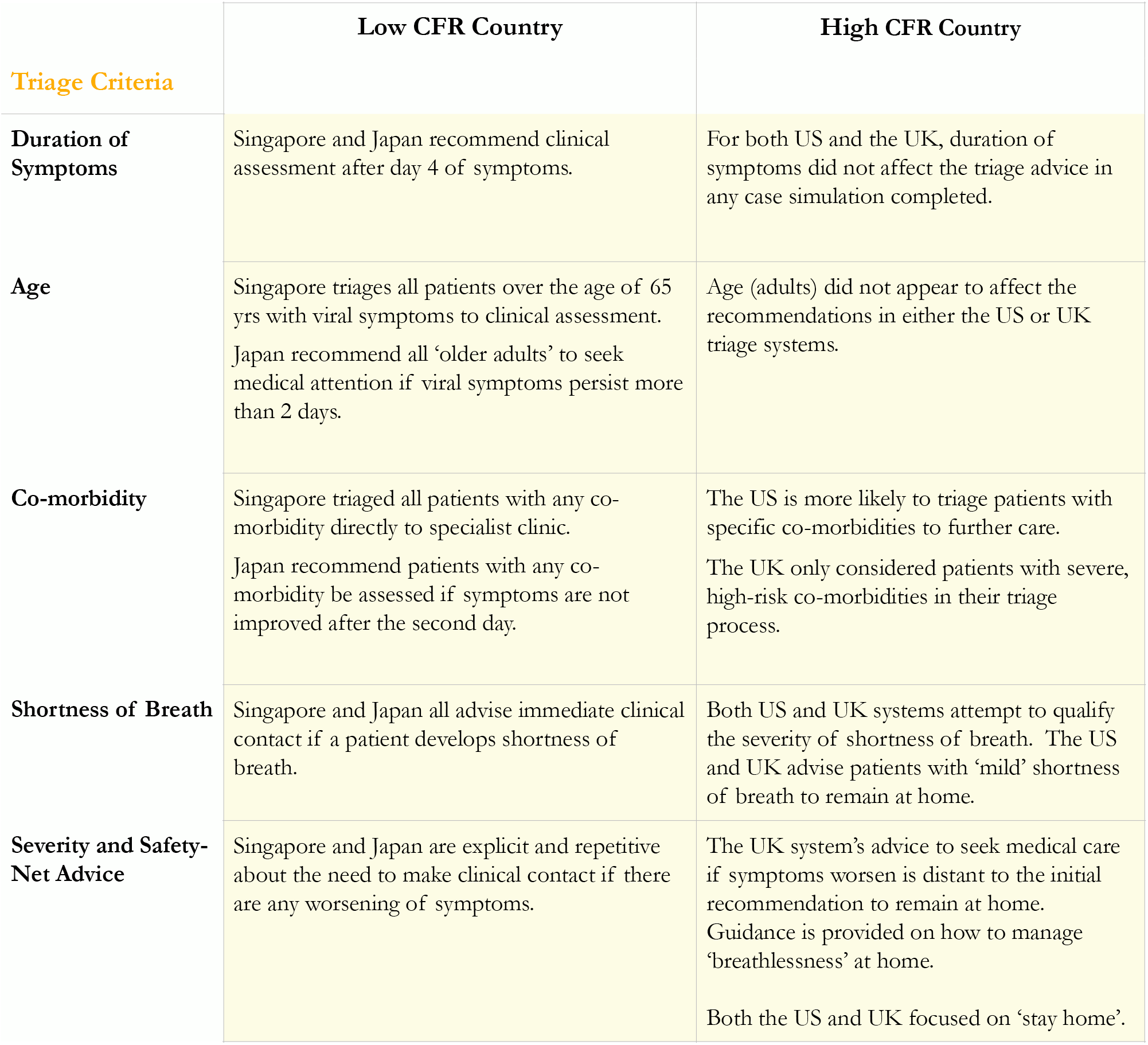
Differences in Triage Criteria between low and high case fatality countries. CFR - Case Fatality Rate; GP - General Practitioner

## DISCUSSION

This case simulation study examined the symptom trackers from four countries. Following application of 52 standardised case simulations to each country’s symptom checker, the percentage of onward referrals were calculated. The low case fatality nations’ (Singapore and Japan) symptom checkers triaged in twice as many cases for direct clinical assessment than the higher case fatality nations (the US and UK). Of great concern was the failure in both the US and UK symptom checkers to triage cases simulating bacterial pneumonia, sepsis and severe COVID-19 on to any healthcare contact.

Symptom checkers are currently being utilised in the pandemic for two purposes: 1) identifying potential cases for testing/surveillance, and 2) identifying ‘unwell’ patients who require medical attention. Both functions are enhanced by the use of symptom checkers when the intention is to ‘catch’ more patients or reach more cases. That is, when symptom checkers are used to identify more cases than would otherwise be detected, and to direct more patients to medical care than would otherwise make healthcare contact, then symptom checkers are merely providing an additional ‘safety-net’, and therefore in such a healthcare support role, the risk of harm from their use is expected to be relatively minimal. Conversely, if symptom checkers are being used to replace the assessment of patients by trained personnel, and are programmed to try and prevent further healthcare contact, then the potential risk of harm for this unproven approach is substantial.

The upside of symptom checkers, particularly during a pandemic is difficult to ignore. By reducing physical patient contacts symptom checkers can potentially save valuable resources, and avoid further viral transmission. Telephone and telemedicine triaging also achieves the protection over further viral transmission, but requires more healthcare staff than symptom checkers. Hence, it is easy to understand the hope of reducing resource expenditure by using symptom trackers as first point of contact.

Evidence to date suggests the majority of cases of COVID-19 resolve after a short, self-limiting viral illness [1]. There are though no discriminatory signs or symptoms [2]. COVID-19 can present like the common cold or flu, or indeed bacterial pneumonia. COVID-19 can also progress quickly [6,35], and can even present with asymptomatic hypoxia [32]. Sifting through the mild colds and self-limiting flus, and trying to determine who will have a mild course of COVID-19, and also trying not to miss bacterial pneumonia, sepsis and signs of COVID-19 pneumonia is quite an ask even for a trained clinician, let alone an automated system.

It is here where Singapore’s symptom checker performs well. The checker is presented on a single webpage, more akin to an online risk-calculator. There are six inputs required from the patient, and one of three outputs generated. The algorithm powering the symptom tracker is not complicated. Age over 65, or the presence of any health condition, or duration of symptoms over four days triggers the advice to seek medical assessment. Any degree of shortness of breath is triaged directly to the Emergency Department. The Singapore symptom checker has ventured into the challenging and unproven area of using symptom checkers to reduce clinical contacts, but has effectively avoided using an algorithm to make clinical decisions. The Singapore Covid-19 Symptom Checker, if utilised by the public, is likely to reduce contacts by the young, fit patients who are early on in the illness, thus off-loading the healthcare burden to some degree, whilst maintaining a relatively low risk to the public.

The UK ‘111’ symptom checker performs poorly in this regard. The algorithm is complex, attempting to quantify symptoms such as shortness of breath, and the overall severity of illness, by asking subjective, qualitative questions with multiple choices. The ‘111’ symptom checker seems to take on a much broader clinical role, and attempts to triage out cases that would normally be triaged in or out on actual clinical assessment. For example, a 72 year old who presents with a seven day history of fever and cough is triaged by the ‘111’ symptom checker to stay at home with no clinical, nursing or healthcare contact. There are few clinicians or nurses that would triage such a patient to stay at home without at least a set of basic observations. The differential in this case includes sepsis, bacterial pneumonia and COVID-19 pneumonia, and whilst it remains possible that fever can persist for seven days in mild/moderate COVID-19, complications or alternative diagnoses are much more likely.

The qualifying questions used by the ‘111’ symptom checker to discriminate between severity will have insufficient discriminatory value in such cases. Further, the wording of the question encourages the self-reporting towards lower categories of illness:

Are you so ill that you’ve stopped doing all of your usual daily activities?

a. Yes - I’ve stopped doing everything I usually do,
b. I feel ill but can do some of my usual activities, or
c. No - I feel well enough to do most of my usual activities.

[Extracted question from ‘111’ Coronavirus Symptom Checker]

It’s the use of absolute and equivocal qualifiers that prevent the severity-qualifying question from achieving any useable clinical triage information: the use of “all” in the question, “everything” in the affirmative answer, and even the negative answer stipulates “most”. Our case simulation demonstrated that answering b), the moderately-severe answer, still triages patients to self-isolate with no healthcare contact. As such, patients with cough and fever for seven days would have to be so severely unwell that they are unable to do anything they usually do to be triaged to any clinical contact.

The UK ‘111 COVID Symptom Checker’ attempts to take on clinical decisions and fails to deliver safe outcomes. Both the ‘111 COVID Symptom Checker’, and the ‘CDC Coronavirus Symptom Checker’ are too aggressive in trying to prevent healthcare contact. Again, beyond the mortality impact, there is no evidence that such an approach actually reduces healthcare burden. Indeed, beyond the established evidence in pneumonia generally [19-22], there is direct evidence that early correction of hypoxia in COVID-19 prevents progression to mechanical ventilation [5], consistent with basic medical principles. Programming symptom checkers to aggressively triage patients to stay home may well increase the burden on intensive care facilities.

Considering that the efficacy of symptom checkers have not been established [17], caution would be advisable. Delay in the correction of hypoxia, failure to commence thromboprophylaxis, and missing the opportunity for earlier initiation of steroids in the hypoxic patient, are all likely to carry a considerable morbidity and mortality cost.

If we are to accept the lesser option of an automated, self-directed triage system over the standard of care offered by the dynamic, experienced clinical assessment, then we must be mindful of what we are asking of the ‘symptom checker’. They are not advanced enough to fulfil the ‘stay home’ intent with any sufficient level of safety. They may though be sufficient enough to assist in the improved identification of at risk patients requiring further clinical assessment, and some form of symptom checker may even be able to contribute to the increased ongoing vigilance required for all patients diagnosed with COVID-19.

### Strengths and Limitations

This case simulation study was conducted using 52 standardised simulated cases. The cases were designed to test specific COVID-19 related scenarios, and as such were symptom-based without the need for subjective interpretation. Nonetheless, there remains a risk of bias, particularly when facing subjective questions. The majority of simulations were though more quantitative, for example duration, age and symptoms, and unlikely to be affected meaningfully by any bias.

The UK data is pooled from all four nations (England, Wales, Scotland and Northern Ireland). England (making up 90% of the total UK population) uses the same ‘111’ COVID-19 patient-led triage system analysed here, whereas Wales, Scotland and Northern Ireland have implemented their own individual patient-led triage systems. It was beyond the scope of this initial investigation to examine each triage system separately. A similar situation applies to the US, where some individual states have implemented their own triage systems.

## CONCLUSION

In this case simulation study, the UK and USA patient-led triage systems (COVID-19 Symptom Checkers) maintained a high disease-severity threshold for onward referral to healthcare assessment. Particular concerns were advising no clinical contact for elderly patients with COVID-19 related symptoms or patients who had developed shortness of breath or any patient with persistent fever. The low case fatality rate countries (Singapore and Japan) utilised symptom checkers to reduce clinical demand whilst maintaining a lower health-risk to patients. Our study suggests, whilst symptom checkers can be of use in the healthcare response to COVID-19, improper use may lead to delayed presentations, and as such an increased healthcare burden, and a likely increased mortality.

## Data Availability

Data is available to reuse and adapt with citation.

## Conflict of Interests

Authors declare no conflicts of interest.

## Ethics Statement

No ethical approval was required for this simulation study. NHS Improvements and NHS Digital were informed of the results of this simulation study on 28/04/20.

## Notes

### Competing Interest Statement

The authors have declared no competing interest.

### Funding Statement

No external funding

### Author Declarations

As this study was a case simulation study with no patient involvement nor involvement of any patient data, the study had no requirement for ethics approval.

